# AI-Generated Clinical Summaries: Errors and Susceptibility to Speech and Speaker Variability

**DOI:** 10.1101/2025.10.29.25339041

**Authors:** Thomas C. Draper, Jason Leake, Timothy Cox, Kathryn Lamb-Riddell, Benjamin E. Johns, John McCormick, Stephen Trowell, Janice Kiely, Richard Luxton

## Abstract

**Summary Box:** *What is already known on this topic:*

- Clinical AI Scribe outputs can contain errors, and the impact of human factors (e.g. communication style, accents, speech impairments) in clinical contexts remains under-characterised.

*What this study adds:*

- In controlled simulations, patient personality and accent did not significantly alter total CAIS errors, with omissions predominating and hallucinations/inaccuracies remaining low.
- Speech-impairment effects were highly varied, with near-perfect recognition for cleft palate and vowel disorders, whereas phonological impairment substantially reduced accuracy.

*How this study might affect research, practice or policy:*

- Supports clinician-in-the-loop deployment with local validation across representative accents and impairment profiles, prioritising detection of clinically critical errors.
- Routine governance should include subgroup performance reporting (accents, impairments) and ongoing audit of error rates.

**Objective:** The study aims to evaluate whether variability in patients’ communication style (personality, international English accents, and speech impairments) affects the accuracy of a Clinical AI Scribe (CAIS), and to identify where performance degrades.

**Method and Analysis:** We conducted simulated primary-care consultations in a purpose-built lab using trained actors. To investigate personality types, four scenarios were enacted, each with five patient-personality types. For accents, human-verified transcripts of consultations were used to generate all doctor/patient combinations of seven different accents (including a synthetic reference voice) across five scenarios. The CAIS produced SOAP-structured summaries that were compared with the transcripts. Errors were classified as omissions, factual inaccuracies, or hallucinations. For speech impairments, public recordings representing five profiles were transcribed and word-recognition accuracy was calculated.

**Results:** Personality types showed no statistically significant differences in errors (all *p*>0.05). Extraversion had the highest total errors (median 3.5), while conscientiousness and agreeableness were lower (1.5 and 2.0, respectively). Across accents, both pairwise tests and group comparisons were non-significant for both patient and doctor voices (patients: *p*=0.851; doctors: *p*=0.98). Omissions predominated, with low rates of hallucinations and factual inaccuracies. Omissions were slightly higher for Chinese- and Indian-accented doctors (both medians 3.0). In contrast, speech impairments differed: cleft palate and vowel disorders were near-perfect, whereas phonological impairment markedly reduced recognition (*p*<0.001).

**Conclusions:** Under controlled conditions, CAIS performance was broadly stable across communication styles and most accents but remained vulnerable to specific speech characteristics, particularly phonological impairment. Future evaluations using real-world, multi-speaker clinical audio are needed to confirm performance.

## Introduction

The application of artificial intelligence (AI) technologies into healthcare systems has rapidly accelerated over recent months, with a particular focus on Ambient Voice Technology (AVT), in particular Clinical AI Scribes (CAISs) [1]. As the adoption of CAISs has become more widespread, so too has the need for independent evidence on their performance and for clear evaluation criteria. In many settings, local implementation expertise is limited, underscoring the importance of unbiased performance data to inform evaluation and rollout [2,3].

These technologies are designed to support clinicians by transcribing spoken interactions into structured clinical summaries, thereby alleviating the administrative burden and potentially improving patient care efficiency [4–7]. However, their deployment in real-world clinical environments introduces complex challenges. Indeed, the presence of errors in the output of CAISs from across the market has been documented, leaving clinician oversight and review crucial to maintaining data integrity and supporting safe clinical decision-making [8–12].

The first stage of CAIS processing is transcribing speech to text. Bias based on gender, age, speech impairment, race, and regional accents has previously been observed in the transcription stage [13]. Transcription errors (caused by human or machine) can have a significant cascading impact on downstream processing [14–16], so whilst it has been shown that Large Language Models (LLMs) can compensate for some missing information from the initial transcript, the masking of key terms propagates thereby reducing accuracy and informativeness of the generated output [17].

Whilst CAIS manufacturers seek to assure users that products have been piloted extensively across several geographical areas with a range of regional accents [18], the literature suggests a lack of corpora that accurately represents the regional variation of spoken English in conversational settings [19]. For example, North American English may result in a higher transcription fidelity compared with British English - typically where the automatic speech recognition (ASR) system has been trained for a North American market, offering secondary markets a less-than-optimal performance [20]. Indeed, non-native English-speaking individuals may experience poorer performance when utilising ASR systems [21,22], with East Asian accents of spoken English experiencing a significantly higher error rate than the North American accent [20]. Indeed, it is documented that errors in early speech recognition technologies have led to actual harm in patients [23].

It has been proposed that the diverse nature of British accents and lack of consistent pronunciation patterns amongst speakers accounts for its less accurate transcription outputs. Notably, it has been observed that British English performs comparably to English spoken with other European accents such as German, Dutch, French, and Polish [20]. With ethnic bias [24] and limited compatibility with speech impairments [13] being acknowledged in ASR, clinicians are compelled to ensure that such patients do not receive an inferior service. However, there is currently insufficient independent research into the impact of accents on the performance of CAISs.

We investigate whether speech and speaker variability (personality, English accents (synthetic and human), and speech impairments [25]) changes the accuracy of a CAIS. Using controlled, standardised consultations, we quantify errors and test for differences by speaker variable and speaker role. We aim to provide empirical evidence on where CAIS performance is robust and where it degrades, to inform pre-deployment testing and subsequent monitoring.

## Method

All recordings were conducted in a simulated primary-care environment at the NHS England South West Centre of Digital Excellence (CoDE), located within the Health Technology Hub at UWE Bristol. Designed and approved by clinicians, this provided controlled room acoustics, realistic spatial configuration, and standardised microphone placement; the same room and hardware were used across conditions. Experiments were conducted between January and September 2025.

### Personality Types

Investigations were performed to examine how the CAIS performed when presented with the same medical scenario, performed by the same individual, but presented in one of five different ways. Four medical scenarios were used (diarrhoea, sleep apnoea, work stress, and headache), each presented with a personality which strongly presented one of the ‘Big Five personality types’ (openness, conscientiousness, extraversion, agreeableness, and neuroticism) [26], leading to a total of 20 recorded consultations, ranging from 7 to 19 minutes in length. Patients were portrayed by professionally-trained actors, who were briefed on both the medical scenario and the required personality. The audio recordings were then directly provided to the CAIS, using the product’s default settings and the Subjective, Objective, Assessment, Plan (SOAP) template, which yielded 20 CAIS generated clinical summaries, each with around 25 to 30 points of information. For each of the 20 encounters, ‘ground-truth’ confirmed-accurate transcripts were also produced.

In order to quantitatively evaluate the generated clinical summary, each was directly marked against its own transcript, and the number of hallucinations, factual inaccuracies, and omissions were counted. In a manner used in some other publications [8,10,27], these are defined as:

- Hallucinations: Instances where the CAIS tool generated content that was not based on the original audio input, causing false information to be included in the summary. E.g. recording “No Known Allergies”, when allergies were never discussed.
- Factual Inaccuracies: Errors where the tool’s summary did not accurately reflect the spoken facts, causing false information to be included in the summary. E.g. recording the wrong name of a medication.
- Omissions: Key pieces of clinical information that were not included in the summary, but were present in the original audio input. E.g. failing to record recent travel history.

Total errors were defined as the sum of omissions, factual inaccuracies, and hallucinations.

### Accents

To standardise lexical content across accents, we generated consultations from human-verified transcripts of the original five actor-recorded consultations. This approach ensured identical wording while varying speaker accent. However, synthesised audio may under-represent natural intonation, colloquialisms, overlap, and turn-taking. Similarly, regional phrases will not be present - for example whilst the script might say “I work from home”, someone with a Scottish dialect may actually say “I work fae hame”.

We used ElevenLabs [28], which provides a commercial service for converting text to speech using a range of different accented voices. Whilst there is a good range of voices, they are generally provided for use in podcasts, AI-generated responses, or multimedia applications, where understandability is important. Hence, whilst the intonation of individual words did vary considerably between the voices that we used, they did not have very heavy accents that might be encountered in primary care.

To address the limitations of synthesised speech, we additionally recorded the five scenarios with humans acting as the patient (English-speaking Irish, Chinese, and Nigerian accents), responding to a synthetic doctor voice reading the clinician lines. Human speakers read the patient script for each scenario to maintain lexical alignment with the synthetic versions. These recordings were processed in the same way as the synthetic voices, enabling direct comparison between human and synthetic patient accents while holding content constant.

It was necessary to generate a strict verbatim transcript that includes disfluencies such as filler words, stutters, and false starts. We used CrisperWhisper [29] to convert the original recording to a reasonably accurate strict transcript, and then hand-edited to improve accuracy. Even so, the strict transcript does not catch some features of the dialogue, such as one person talking over the other. We edited it into a form where the speaker was identified so that a small Python program could use ElevenLabs to generate a dialogue consisting of alternated speech by each participant. At the end of this process, we had a set of recordings mirroring the original consultation recording, but with different voices playing each role in each recording.

Because all speakers have an accent, there is no true “baseline” accent. Therefore, for each consultation, we generated a new audio file with the doctor as a clear articulate, computer-generated ‘synthetic’ voice. This ‘synthetic’ voice was used as our baseline voice as we felt it would be the easiest for the scribe to understand. The patient was an accented voice. We also generated the same consultation with the doctor as an accented voice, and the patient as the ‘synthetic’ voice. The strength of the accents varied from light-to-moderate-to-strong.

Five different scenarios were used for each combination: diarrhoea, headache, prostate, skin rash, and sleep apnoea, and we used the CAIS to generate a clinical summary for each accent combination for each scenario. Five accents (American, Chinese, Indian, Irish, Scottish) × two voice roles (doctor accented/patient synthetic; patient accented/doctor synthetic) × five scenarios = 50 consultations, plus five synthetic-synthetic baselines, plus 15 human recordings (5 scenarios, human patient accented/doctor synthetic), totalling 70 consultations and generated clinical summaries. Errors were then determined by comparing each AI-generated clinical summary to the human-verified transcript.

### Speech Impairments

In order to investigate the CAIS’s ability to comprehend voices with speech impairments, we obtained publicly available audio recordings of speakers exhibiting five distinct speech-impairment profiles (phonological impairment, vowel disorder, childhood apraxia of speech, articulation disorder, and cleft palate) [30,31]. To approximate a clinical dictation scenario, each recording was paired with a ‘ground-truth’ (human-confirmed) transcript prepared in advance. Each audio file was presented to the CAIS, which then attempted to create a transcript (summarisation was not attempted, as these were not medical conversations).

From each transcript we computed the proportion of words correctly recognised by aligning the CAIS output to the ground-truth transcript, and calculating the percentage of exact matches. For each impairment category, the outcome was the percentage of words correctly recognised, expressed as a score between 0% and 100%.

### Statistical Analysis

Statistical analyses, using non-parametric tests, of these results were conducted to evaluate the significance of observed variations. Statistical analysis was conducted using R in R Studio [32,33]. Non-parametric analysis was used to compare number of errors and error rates, employing the Wilcoxon signed-rank test, Mann-Whitney U test, and the Kruskal-Wallis test. Medians are reported along with the interquartile range (IQR) in brackets immediately following. Significance is taken as *p* < 0.05.

### Patient and Public Involvement Statement

A Public and Patient Involvement (PPI) group (Citizens Reference Group) contributed to study design and reviewed preliminary findings, providing feedback that informed subsequent stages of the research.

## Results

### Personality Types

To investigate whether patient personality traits influence the accuracy of CAIS outputs, a controlled study was conducted using simulated consultations. Four clinical scenarios (diarrhoea, sleep apnoea, work stress, and headache) were enacted by trained actors, each performed in alignment with one of five personality types derived from the Big Five model: agreeableness, conscientiousness, extraversion, neuroticism, and openness [26]. This design yielded a total of 20 consultations, allowing for cross-comparison of CAIS performance across both scenario type and patient communication style.

As shown in figure 1, all personality types were associated with some degree of error in the CAIS-generated clinical summaries, including hallucinations, factual inaccuracies, and omissions. While there was noticeable variation in the error distribution across personality types, one-sample Wilcoxon signed-rank tests were performed to assess whether the total number of errors for each personality type differed significantly from zero (i.e. whether any meaningful increase in errors was introduced), and none of the differences reached statistical significance (all being *p* > 0.05, full values in table S1, supplementary information). This suggests that, from a quantitative standpoint, the CAIS responded similarly across the spectrum of investigated patient communication styles.

**Figure 1:**
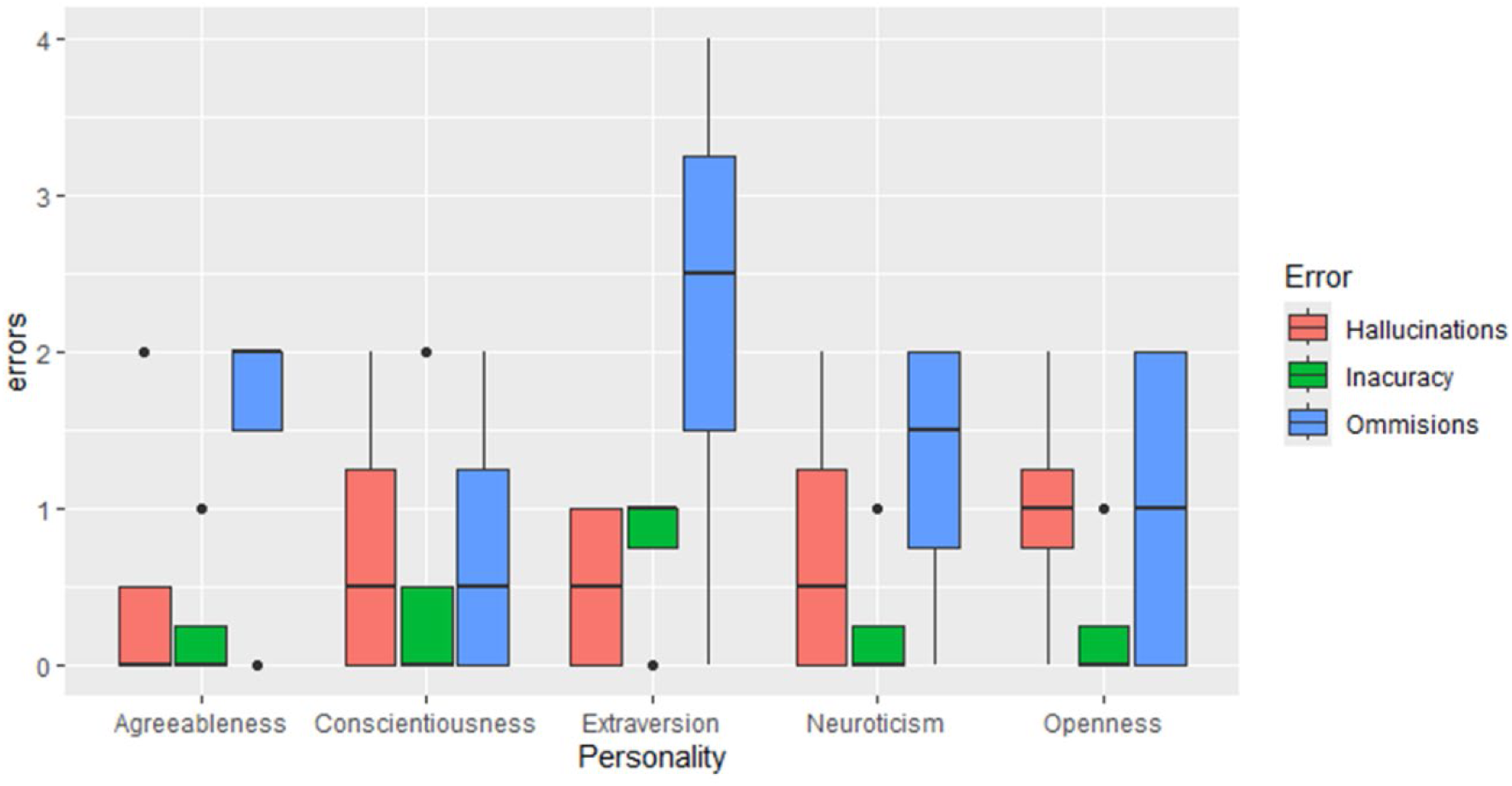
Distribution of hallucinations, factual inaccuracies, and omissions in CAIS-generated clinical summaries across five patient personality personas. Extraversion showed the highest median total errors, primarily due to omissions, while conscientiousness and agreeableness had slightly lower error rates. Differences between personality types were not statistically significant (all p > 0.05).

Among the personality types, extraversion produced the highest number of total errors (median of 3.5), driven primarily by omissions. This aligns with expectations, as extraverted speech can be more dynamic, fast-paced, and tangential, potentially challenging for speech-to-text and summarisation algorithms. However, the lack of statistical significance suggests that this apparent trend may be attributable to random variation rather than a systemic limitation.

Conscientious and agreeable personas, often associated with more cooperative communication styles, showed slightly lower overall error rates (medians of 1.5 and 2.0, respectively), and exhibited fewer hallucinations and inaccuracies. Openness and neuroticism showed more variability in the type and frequency of errors, particularly in emotionally complex consultations (e.g. work stress), yet still did not reach statistical significance.

### Accents

Analysis of the CAIS-generated clinical summaries (shown in figure 2) revealed no statistically significant differences in error rates associated with either patient (Kruskal-Wallis, *p* = 0.851) or doctor accents (Kruskal-Wallis, *p* = 0.980); see figure S1 and tables S2 and S3 in the supplementary information. Pairwise Wilcoxon signed-rank tests were also conducted to compare all accent combinations within both patient and doctor voice conditions. None of the pairwise comparisons reached statistical significance, reinforcing the conclusion that CAIS error rates do not differ meaningfully between any specific accent pairings. Full *p*-values are provided in tables S4 and S5 (both supplementary information).

**Figure 2:**
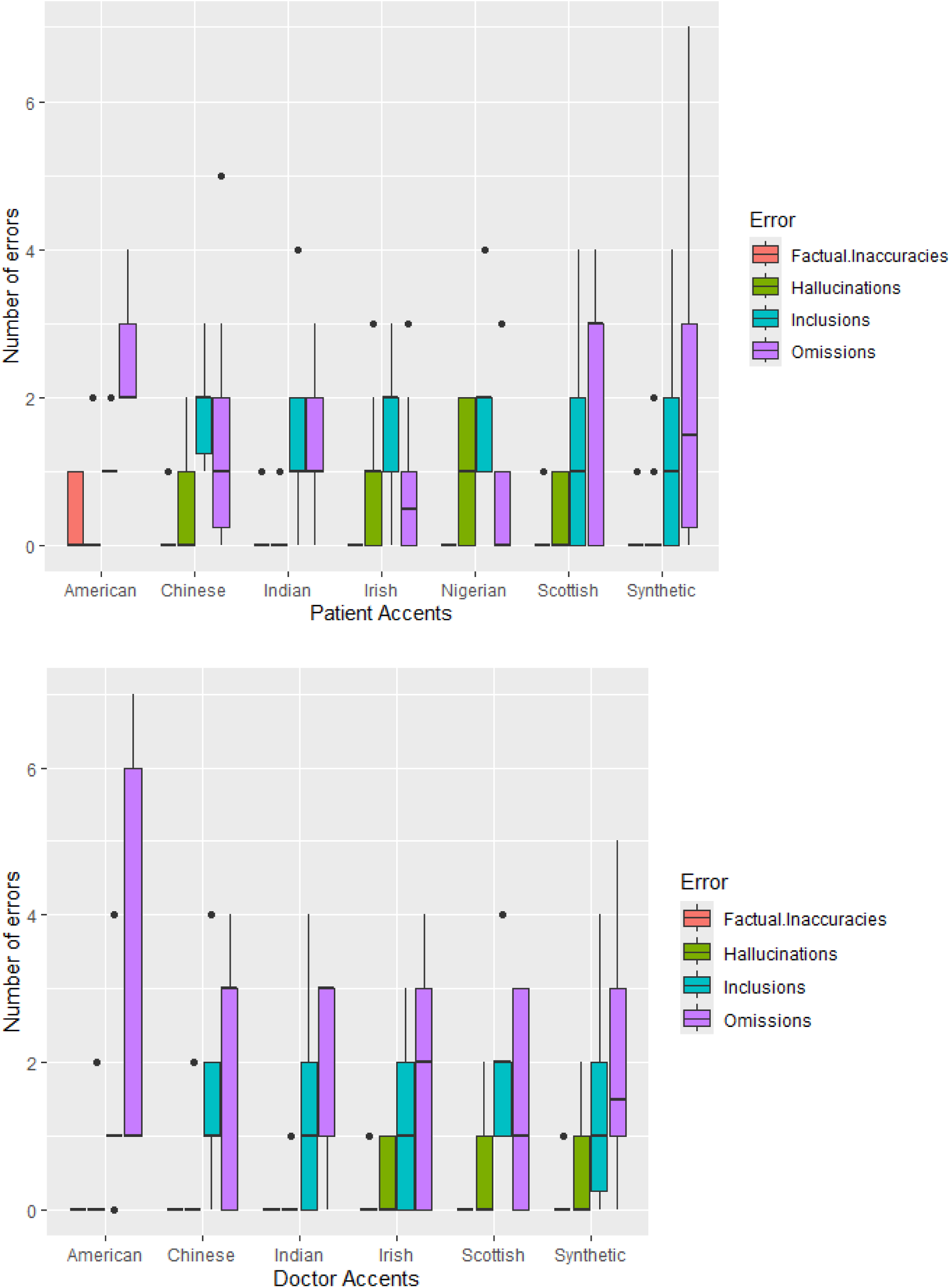
Errors for (a) patient, and (b) doctor, accents compared with the audio transcript, as a boxplot. No significant differences were observed across the accents. The American-accented doctor voice showed a modest increase in omissions, although this was not statistically significant. Each box represents the aggregate error count (hallucinations, inaccuracies, omissions) per condition.

Applying the accent to the doctor versus the patient component made no appreciable difference to total error counts (Mann-Whitney U test, *p* = 0.4654). This pattern was consistent across scenarios, with omissions predominating and hallucinations/factual inaccuracies remaining low in both role assignments. Therefore, the CAIS appeared role-insensitive to accent variation: whether the accent originated from the clinician or the patient did not materially alter performance.

Despite the lack of statistical significance, some nuanced patterns were observed. Among patient accents, no individual accent emerged as clearly more error-prone, although Scottish-accented patients exhibited the highest median omission rate (median = 3.0), compared to other accents which typically ranged between 1.0 and 2.0.

For doctor accents, Chinese and Indian accents showed the highest median omissions (both at 3.0). The American-accented doctor did not exhibit the highest median (1.0), but showed a markedly wider distribution, with omissions reaching an upper quartile value of 6.0. This suggests that while overall performance was similar, occasional consultations exhibited more pronounced information loss. These findings, though not statistically significant, indicate that some accent-voice pairings may pose greater challenges to CAIS performance than others.

Total errors did not significantly differ across human and synthetic patient accents (Kruskal-Wallis, *p* = 0.961), indicating comparable overall CAIS performance and supporting the use of high-quality synthetic voices as a reasonable proxy for accent testing. This can be seen in figure 3. At the error-type level, there were small distributional differences between some error types (hallucinations and omissions), which also varied by scenario. Because scenario effects were not the focus of this study, the aggregated scenarios are compared here, and a per-scenario breakdown can be seen in figure S2 in the supplementary information, for the interested reader.

**Figure 3:**
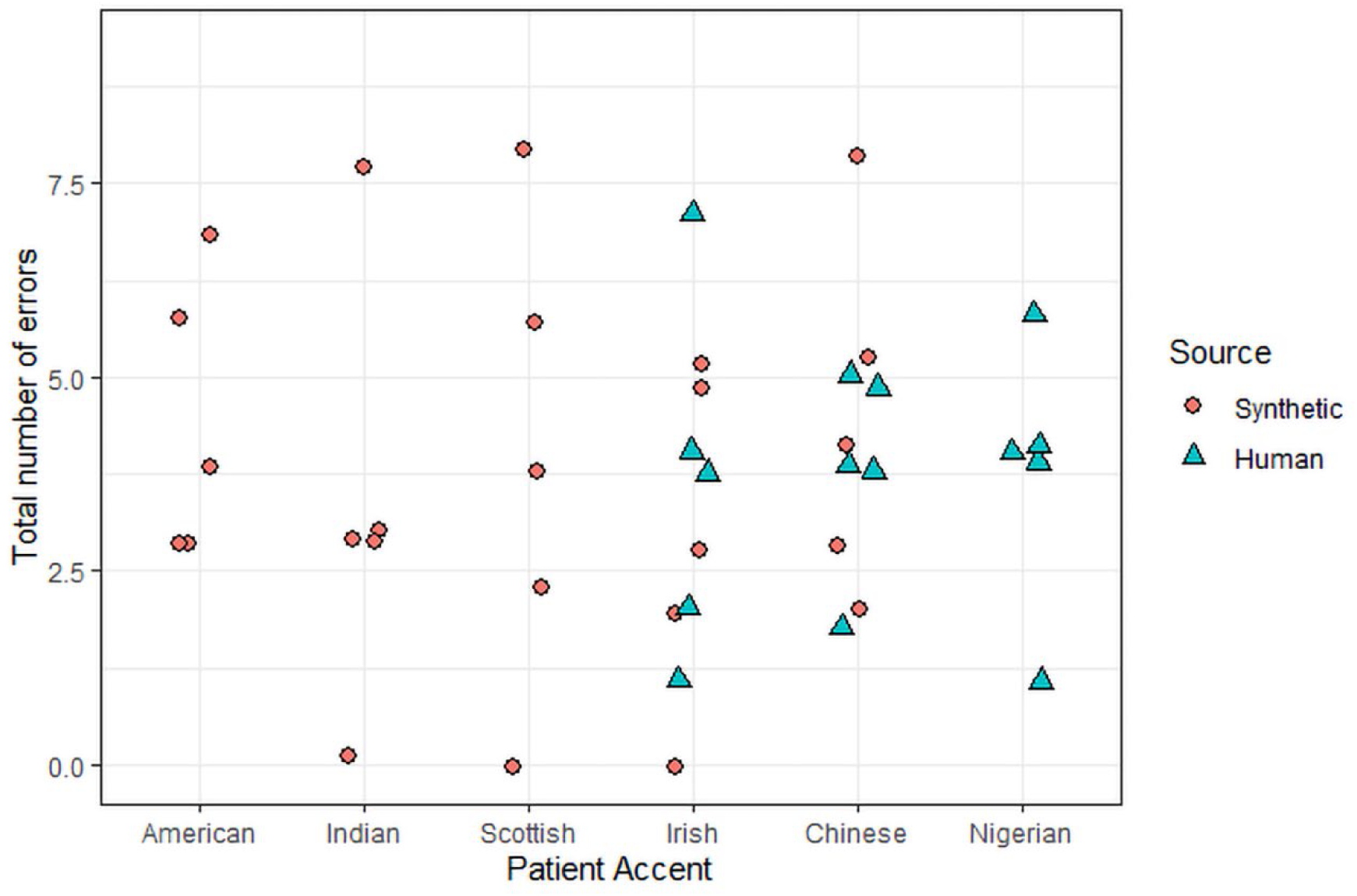
Total CAIS errors by patient accent and source (synthetic vs human). Each point represents one consultation; circles denote synthetic speech and triangles human speech for the patient. Across accents, total error counts were similar for human and synthetic voices (Kruskal-Wallis, p = 0.961). Scenario-level details can be seen in figure S2 in the supplementary information.

Factual inaccuracies and hallucinations were minimal across all accent conditions, with medians at or near zero, further reinforcing the conclusion that most performance variability arises from omission-type errors.

### Speech Impairments

Recordings of individuals with five types of speech impairments [30,31] were processed by the CAIS, and its transcripts were evaluated by comparing the percentage of correctly recognised words against ground truth transcripts. Figure 4 summarises the distribution of recognition accuracy (100% being completely accurate, and 0% being completely inaccurate) for each impairment type. Notably, some speech impairment audio recordings were difficult for the authors to understand themselves. The speech impairments assessed were phonological impairment, vowel disorder, childhood apraxia of speech, articulation disorder, and cleft palate.

**Figure 4:**
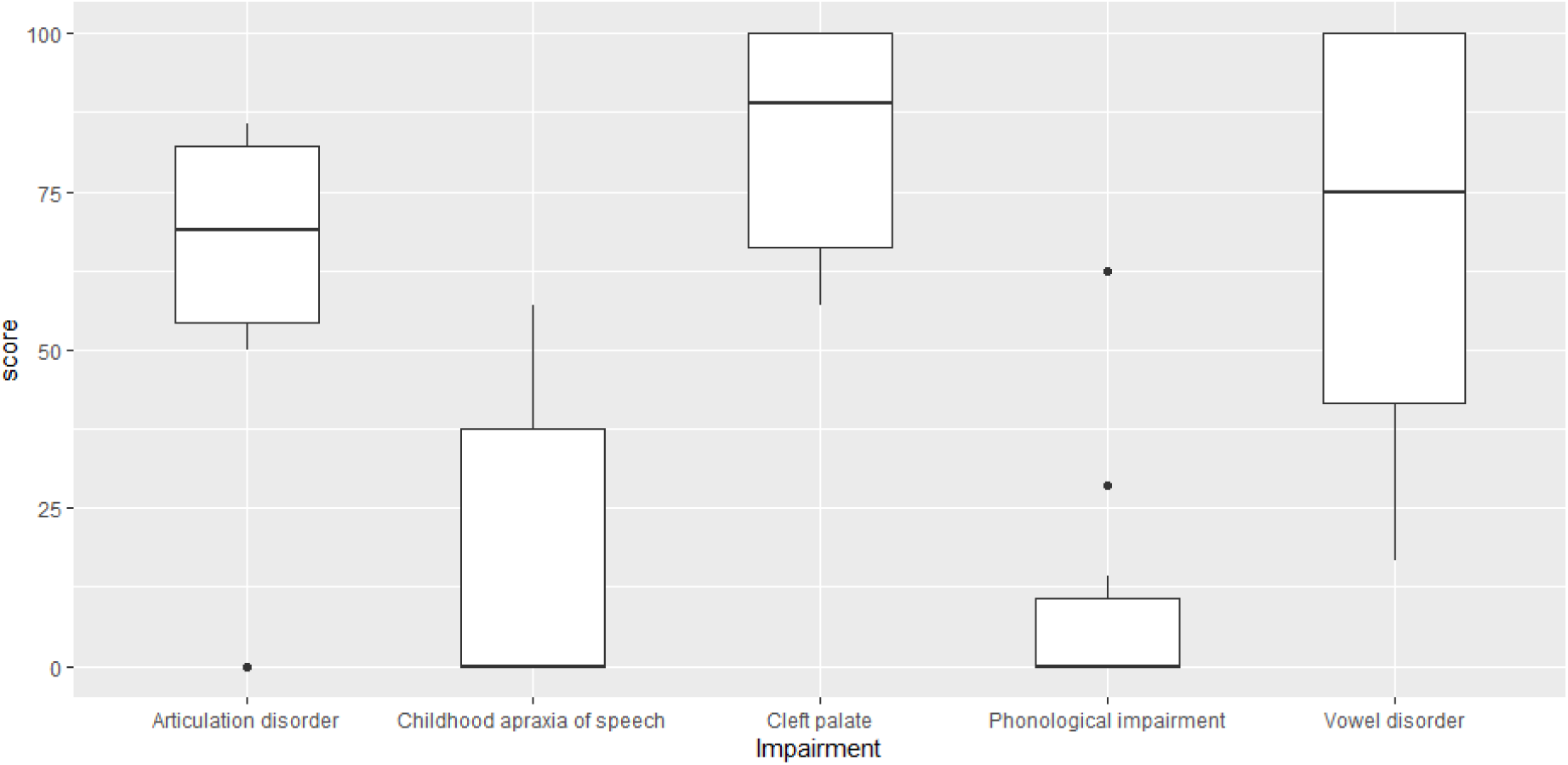
A box and whisker plot of the accuracy of the CAIS at recognising different speech affected by various speech impairments. The score is the percentage of words that are correctly recognised by the CAIS.

A one-sample Wilcoxon test was performed to assess the statistical significance of the deviation from 100% speech recognition. Cleft-palate recordings had the highest recognition accuracy and did not differ significantly from perfect recognition (one-sample Wilcoxon, *p* = 0.181); whilst the vowel-disorder recordings showed wider variability but were also not significantly different from 100% (*p* = 0.371). In contrast, phonological impairment had a significant reduction in accuracy (*p* < 0.001). Overall, these results demonstrate that AI transcription accuracy varies markedly by type of speech impairment, with cleft palate and vowel disorders showing relatively preserved recognition, while phonological and articulation deficits lead to substantial transcription errors. All *p*-values can be found in table S6 in the supplementary information.

## Discussion

### Speech and Speaker Variability

This study explored how different aspects of human communication (ranging from interpersonal style and accent to the presence of speech impairments) affect the performance of a CAIS. Across all these dimensions, the system displayed a broadly stable performance, with no statistically significant differences in overall error rates. Nevertheless, the patterns observed provide important insights into where variability does, and does not, appear to challenge CAIS transcription accuracy.

Within the controlled simulation framework, trained actors portrayed standardised medical scenarios using distinct personality types derived from the Big Five model [26]. While the system did record some transcription errors across all scenarios and personality types, no statistically significant differences were observed between groups. These findings contribute an important piece of evidence to the growing field of AI integration in clinical settings, suggesting that such systems may be broadly resilient to interpersonal variability in patient communication styles.

It is noteworthy that extraverted personas were associated with the highest overall error rates, particularly omissions. Extraverted communication is often characterised by both high verbal output, and spontaneous topic and energy shifts, which can result in reduced structural clarity [34–36]. Although these are factors that could potentially challenge natural language processing (NLP) systems, the absence of statistically significant variation indicates that the CAIS does not exhibit systematic bias in response to specific personality traits. This is a reassuring finding, indicating that the system’s output quality is not disproportionately affected by differences in patient communication style. By contrast, conscientious and agreeable personas showed slightly fewer errors, potentially reflecting their generally more organised and cooperative communication patterns [37,38], which could align more closely with the input expectations of CAIS models. However, this should not be interpreted as strong evidence of performance improvement, given the lack of statistical significance. It does, however, raise interesting hypotheses for future studies: namely, whether communication style training or calibration could improve CAIS performance.

The analysis examining the influence of speaker accents suggested broad robustness under the study conditions, with no statistically significant differences in total error rates across examined international English accents. While the original design relied on synthesised accents, the added human-accent replication (Irish, Chinese, Nigerian) showed no statistically significant difference in total errors relative to synthetic accents, suggesting that well-generated synthetic voices can approximate accent effects for CAIS evaluation. Nonetheless, error-type distributions (particularly omissions and hallucinations) showed modest scenario-specific shifts, warranting caution when interpreting results from synthetic speech alone.

The slightly elevated omissions observed in the American-accented doctor voice, particularly the wider variability and higher upper limits, further suggest that even subtle vocal features may influence how CAIS systems process information from dialogue. Similarly, higher median omissions in Chinese and Indian accented doctor voices, and in Scottish accented patients, point to real-world relevance that should be explored further. While these patterns fall within normal variability, they may still hold practical relevance in real-world use, where naturally accented speech includes more marked prosody, region-specific phrasing, and spontaneous interactional features that are not always fully captured by synthesised speech.

The analysis of speech impairments revealed more pronounced performance variability. Speakers with cleft palate and vowel disorders did not differ, statistically, from perfect accuracy. In contrast, phonological impairments produced a dramatic collapse in recognition (*p* < 0.001), indicating that the system had extreme difficultly extracting accurate information. These discrepancies have direct implications for deploying CAISs in clinical contexts: while models may perform adequately for patients with some impairments, they are entirely unreliable for those whose speech patterns are more complex [39].

In general, the combined evidence indicates that CAIS performance is broadly stable across personality-driven communication styles and most accent conditions tested; however specific speech impairments can substantially degrade performance. Importantly, these findings also demonstrate that the persistent baseline error rate, as previously reported for various CAISs [8,10,12,40], was observed across the tested personality types and accents. This underscores the ongoing need for robust human oversight in the deployment of such technologies, and highlights that further optimisation of these systems remain warranted [41].

Beyond these empirical findings, this work makes a methodological contribution by combining personality theory with AI performance assessment. To our knowledge, it is the first evaluation of a clinical AI explicitly structured around variability in the user’s (patient’s) personality, an approach that could be extended to other human-AI interfaces. As generative and summarising AI tools (such as CAISs) become more prevalent in healthcare workflows, understanding how they interact with human variability will be crucial not only for safety and accuracy, but also for trust and adoption by clinicians and patients alike.

### Implications for Clinical Practice

These findings have immediate implications for the safe and effective deployment of CAISs across primary, community, and acute settings. The overall stability of performance across patient communication styles and international English accents, combined with a persistent baseline error profile dominated by omissions, indicates that CAISs should be implemented as “human-in-the-loop” systems with explicit verification of key fields (including any codes [42]) before note completion. In particular, presenting complaint, salient findings, working diagnosis, medicines and allergies, decisions (tests, prescriptions, referrals), safety-netting, and agreed care plan should be subject to routine clinician verification at sign-off.

The pronounced degradation observed for specific speech-impairment profiles underscores the need for predefined “switch-off” criteria and reasonable adjustments. Organisations and/or Health Authorities should articulate conditions in which CAIS use is contraindicated (e.g., severe phonological impairment) and ensure accessible alternative documentation routes. Recording the reason for non-use will help avoid inequities and support audit.

Operationalisation should sit squarely within already established clinical risk-management practice, with quality assurance at two levels: (i) set-up validation (room acoustics, microphone placement, device/network checks) and (ii) ongoing assurance (random note audit, incident capture, and drift monitoring of error rates with pre-specified thresholds and escalation paths [43]). Prior empirical work on acoustic and informational noise can inform technical minima (e.g., approved microphone types and placement, background-noise tolerances, and pre-clinic audio checks [10]).

Accountability must remain clear: irrespective of team composition, the clinician of record is responsible for the final note. Where multi-professional pathways or training environments are involved, a named reviewer model within the same session should be the default, with any deferral governed by local policy and audit. Given the broadly similar performance across personalities and accents under study conditions, these governance measures should not be selectively applied to particular patient groups, thus supporting fairness and consistency.

### Limitations and Future Directions

The use of trained actors in a controlled environment allowed for experimental consistency, but it also introduces limitations. Real-world consultations are often more variable and emotionally nuanced than even the best simulations can capture. In routine practice, consultations frequently involve overlapping talk, brief interruptions, and contributions from third parties (e.g. a carer), which increase the risk of speaker misattribution and missed content (omissions). Personality traits may also co-occur with other factors (e.g. age, stress, language proficiency) that further affect communication style and, consequently, CAIS performance. Future research should explore these real-world dynamics using large and diverse datasets with multiple CAISs, to investigate whether different systems exhibit similar error profiles under comparable conditions.

The main limitation of the accent analysis is the partial use of synthesised speech. While the use of synthetic voices standardises lexical content, it can lack natural conversational dynamics, region-specific phrasing, and prosody, which could hide accent-related effects on CAIS performance. Although human-accent recordings were also utilised, these were ‘read-speech’ [44] with a synthetic clinician voice, so spontaneous overlap, pauses, and third-party interjections were absent. Prospective studies should use naturalistic, multi-speaker consultations with naturally accented speech to confirm CAIS reliability and to detect any clinically relevant performance differences.

The speech impairments analysis necessarily relied on scripted, non-medical recordings, which lack the spontaneous vocabulary, background noise, and emotions typical of real consultations. To enhance clinical applicability, future work should involve collecting consented medical speech from individuals with diverse speech impairments.

### Conclusion

Across personality-driven communication styles, international English accents, and a range of speech-impairment profiles, the CAIS demonstrated broadly stable performance. Personality traits were not associated with statistically significant differences in total errors, and accent effects were similarly non-significant. The majority of variability arose from omissions, with counts of hallucinations and factual inaccuracies remaining low. In contrast, phonological impairments produced a marked collapse in recognition accuracy, whereas cleft palate and vowel disorders were comparatively well recognised.

Collectively, these findings indicate that current CAIS technology is largely resilient to everyday interpersonal and linguistic variability, yet remains vulnerable to specific speech characteristics that can impair transcription accuracy. Importantly, performance did not appear to be disproportionately influenced by communication style, a critical consideration for fairness, reliability, and equity in patient care.

## Acknowledgements

The authors particularly thank Caroline Hadley and the UWE Drama and Acting school for their skills portraying the various personality types, as well as to our voice-actors Ibidapo Williams, Alex Yue, and Rónán Tynan. We also thank NHS England South West for supporting the establishment of the CoDE at UWE Bristol, and the participants of the Citizens Reference Group for their contributions.

## Contributions

JMC, ST, JK, and RL conceived the study. TCD, TC, JK, and RL designed the study. TCD, JL, TC and KLR were involved in the acquisition of data. TCD, JL, TC, and RL analysed the data. TCD, JL, JMC and BEJ drafted the manuscript. All authors approved the final version of the manuscript. The corresponding author (TCD) is the guarantor. The corresponding author attests that all listed authors meet authorship criteria and that no others meeting the criteria have been omitted.

## Ethical Approval

Ethical approval for the work undertaken was granted by the University of the West of England Ethics committee – No: CHSS.24.05.199.

## Data Availability Statement

Outputs from the commercial Clinical AI Scribe, along with transcripts of the original audio files, will be made available upon reasonable request to the corresponding author.

## Transparency Statement

The corresponding author affirms that the manuscript is an honest, accurate, and transparent account of the study being reported; that no important aspects of the study have been omitted; and that any discrepancies from the study as planned have been explained.

