## Supplementary Information for "AI-Generated Clinical Summaries: Errors and Susceptibility to Speech and Speaker Variability"

(b) NHS England South West, South West House, Blackbrook Ave, Taunton, TA1 2PX, UK


### Supplementary Information

#### Tables

***Table S1:*** *The results of a one-sample Wilcoxon signed-rank test, indicating if the total number of errors in the AI-generated clinical summary associated with the personality type has a significant difference from zero. All personality types had p > 0.05, indicating that there is no significant difference.*

| Personality Type | p Value |
|---|---|
| Agreeableness | 0.0890 |
| Conscientiousness | 0.0975 |
| Extraversion | 0.1250 |
| Neuroticism | 0.1814 |
| Openness | 0.0975 |

***Table S2:*** *Summary of error counts across patient accents, aggregated over five clinical scenarios, with human and synthetic voices. Median values are reported for omissions, factual inaccuracies, and hallucinations, with interquartile ranges (IQRs) shown in brackets. Although no statistically significant differences were observed, Scottish and American accented patients exhibited numerically higher median omissions, and Irish-accented patients had slightly more hallucinations.*

| Patient Accent | Omissions | Factual Inaccuracies | Hallucinations |
|---|---|---|---|
| American | 2.0 (2–3) | 0.0 (0–1) | 0.0 (0–0) |
| Chinese | 1.0 (0.25–2) | 0.0 (0–0) | 0.0 (0–1) |
| Indian | 1.0 (1–2) | 0.0 (0–0) | 0.0 (0–0) |
| Irish | 0.5 (0–1) | 0.0 (0–0) | 1.0 (0–1) |
| Nigerian | 0 (0–1) | 0.0 (0–0) | 1.0 (0–2) |
| Scottish | 3.0 (0–3) | 0.0 (0–0) | 0.0 (0–1) |
| Synthetic | 1.5 (0.25–3) | 0.0 (0–0) | 0.0 (0–0) |

***Table S3:*** *Summary of error counts across doctor accents, aggregated over five clinical scenarios. Median values and IQRs are shown for each error type. Although differences were not statistically significant, Chinese and Indian-accented doctors were associated with the highest median omissions, and the American-accented doctor voice demonstrated a broader IQR for omissions, indicating occasional elevated error rates.*

| Doctor Accent | Omissions | Factual Inaccuracies | Hallucinations |
|---|---|---|---|
| American | 1.0 (1–6) | 0.0 (0–0) | 0.0 (0–0) |
| Chinese | 3.0 (0–3) | 0.0 (0–0) | 0.0 (0–0) |
| Indian | 3.0 (1–3) | 0.0 (0–0) | 0.0 (0–0) |
| Irish | 2.0 (0–3) | 0.0 (0–0) | 0.0 (0–1) |
| Scottish | 1.0 (0–3) | 0.0 (0–0) | 0.0 (0–1) |
| Synthetic | 1.5 (1–3) | 0.0 (0–0) | 0.0 (0–1) |

***Table S4:*** *Two-sided Wilcoxon signed-rank tests comparing error rates between all patient accent pairings. No comparison reached statistical significance.*

| Accent 1 | Accent 2 | *p*-value |
|---|---|---|
| American | Chinese | 0.803 |
| American | Indian | 0.373 |
| American | Irish | 0.322 |
| American | Nigerian | 0.828 |
| American | Scottish | 0.833 |
| American | Synthetic | 0.271 |
| Chinese | Indian | 0.350 |
| Chinese | Irish | 0.355 |
| Chinese | Nigerian | 0.799 |
| Chinese | Scottish | 0.950 |
| Chinese | Synthetic | 0.267 |
| Indian | Irish | 0.951 |
| Indian | Nigerian | 0.392 |
| Indian | Scottish | 0.831 |
| Indian | Synthetic | 0.905 |
| Irish | Nigerian | 0.708 |
| Irish | Scottish | 0.711 |
| Irish | Synthetic | 0.837 |
| Nigerian | Scottish | 1.000 |
| Nigerian | Synthetic | 0.633 |
| Scottish | Synthetic | 0.886 |

***Table S5:*** *Two-sided Wilcoxon signed-rank tests comparing error rates between all doctor accent pairings. No comparison reached statistical significance.*

| **Accent 1** | **Accent 2** | ***p*-value** |
|---|---|---|
| American | Chinese | 0.671 |
| American | Indian | 0.750 |
| American | Irish | 0.589 |
| American | Scottish | 0.672 |
| American | Synthetic | 0.404 |
| Chinese | Indian | 0.750 |
| Chinese | Irish | 1.000 |
| Chinese | Scottish | 1.000 |
| Chinese | Synthetic | 0.905 |
| Indian | Irish | 1.000 |
| Indian | Scottish | 1.000 |
| Indian | Synthetic | 0.866 |
| Irish | Scottish | 1.000 |
| Irish | Synthetic | 0.886 |
| Scottish | Synthetic | 1.000 |

***Table S6:*** *The results of a one-sample Wilcoxon signed-rank test, to assess the significance of the deviation from 100% speech recognition in the CAIS transcript. Articulation disorders and phonological impairment both had a statistically significant difference, with* $p < 0.05$.

| **Speech Impairment** | ***p*-Value** |
|---|---|
| Articulation disorder | 0.0355 |
| Childhood apraxia | 0.0545 |
| Cleft palate | 0.1814 |
| Phonological impairment | 0.0006 |
| Vowel disorder | 0.3711 |

#### Figures

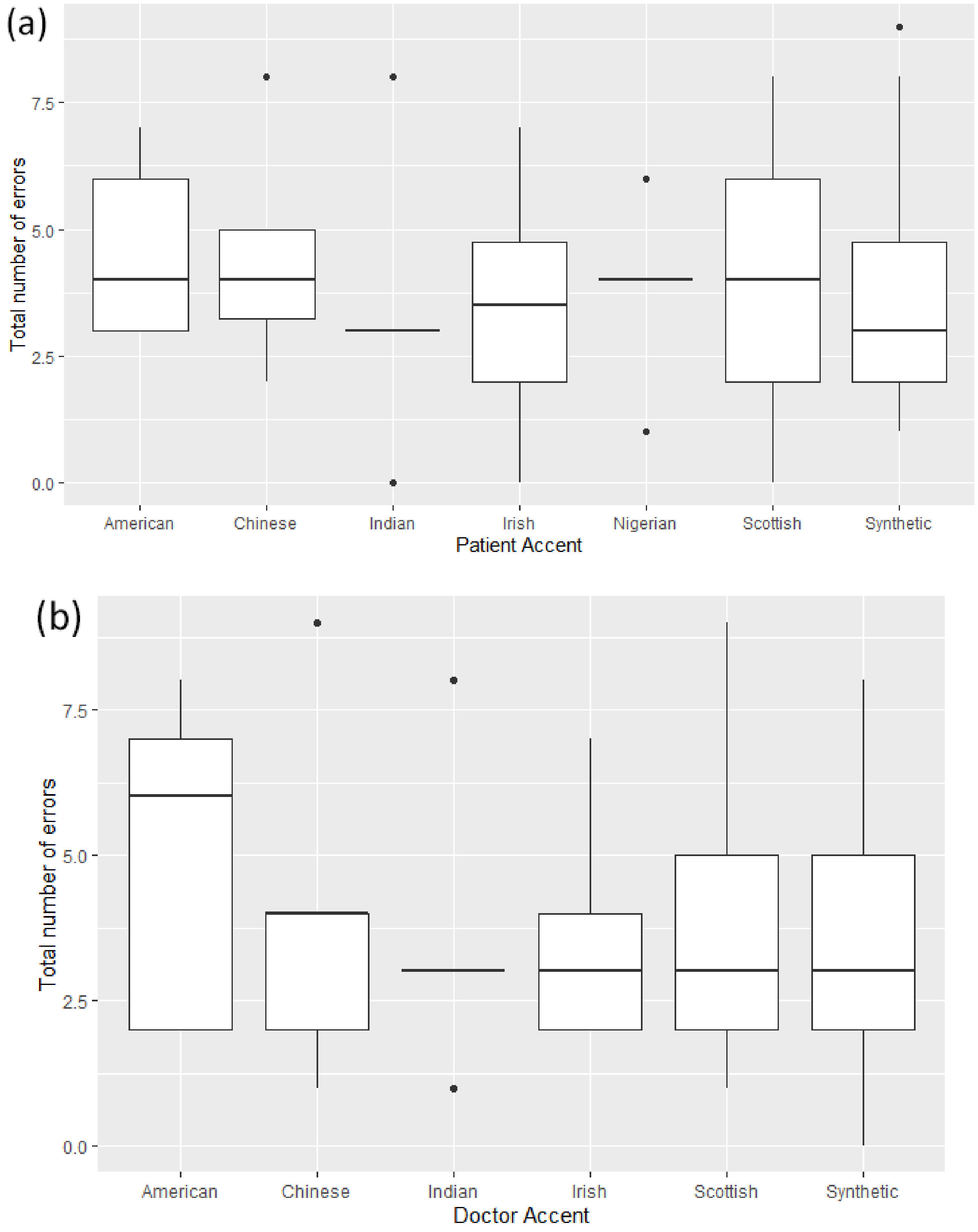


***Figure S1:*** *Comparison of Clinical AI Scribe (CAIS) performance when accents are applied to either the patient or the doctor component of the consultation. (a) Median total errors per consultation with a synthetic doctor and accented patient; Kruskal-Wallis, p = 0.851 (no between-accent differences). (b) Synthetic patient and accented doctor; Kruskal-Wallis, p = 0.980 (no between-accent differences). A direct comparison of doctor- vs patient-assigned accents showed no difference in totals (Mann-Whitney U, p = 0.4654; see main text).*

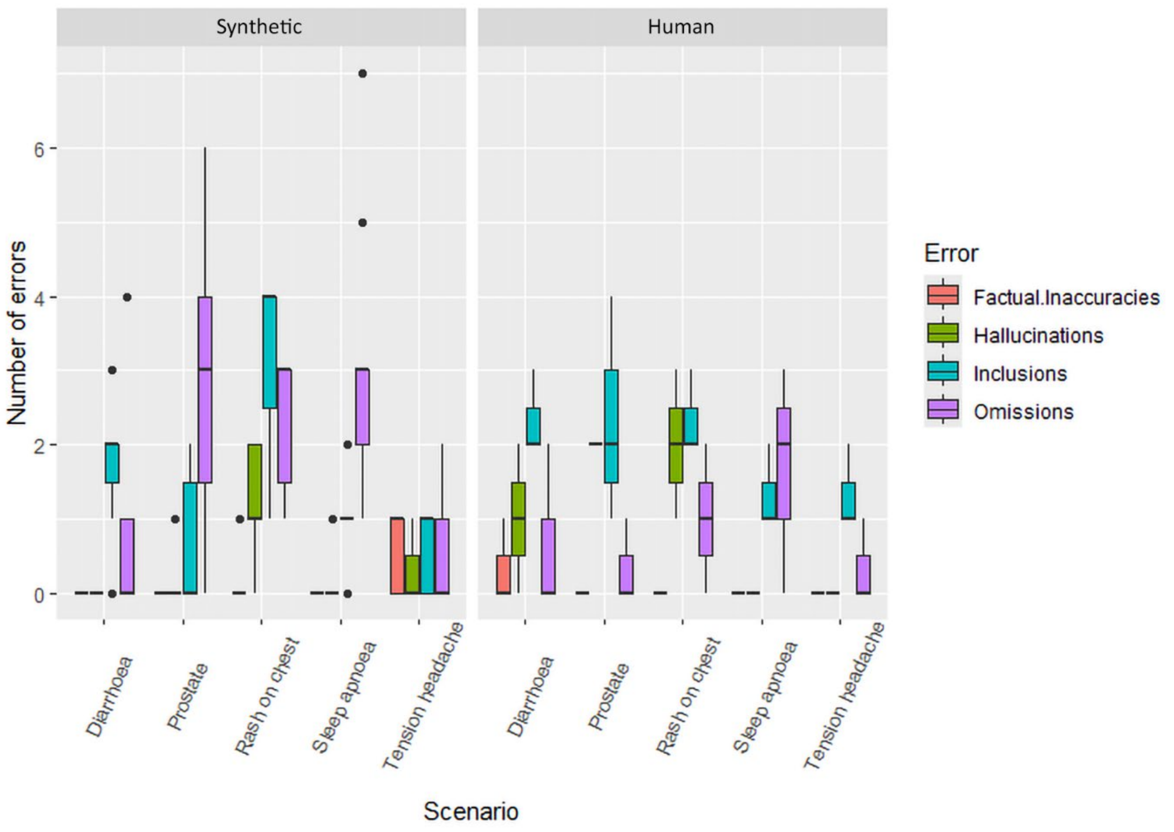


***Figure S2:*** *Distribution of error types across individual clinical scenarios for each accent condition. Each box represents the aggregated error count for a given scenario (diarrhoea, prostate, rash, sleep apnoea and headache) under different accent pairings. The left-hand side collates computer-synthesised voices, and the right-hand side collates human voices. Although total error counts were similar across accents, scenario-level patterns show minor variation in the relative proportion of omission versus hallucination errors.*
